# Associations between work characteristics and large joint osteoarthritis: a cross-sectional study of 285,947 UK Biobank participants

**DOI:** 10.1101/2024.08.05.24311461

**Authors:** A Hashmi, S Scott, M Jung, Q-J Meng, JH Tobias, RA Beynon, BG Faber

## Abstract

**Objectives:** Shift work-induced circadian rhythm disruption has been identified as a risk factor for specific diseases. Additionally, physically demanding work has been linked to osteoarthritis. This study investigated the independent associations of shift work and physical work with risk of large joint osteoarthritis.

**Design:** UK Biobank participants completed questionnaires detailing their employment status, including shift work, night shifts, heavy manual work and prolonged non-sedentary work. Responses were categorised into binary and categorical variables. Knee and hip osteoarthritis diagnoses were extracted from hospital records and osteoarthritis (any site) was self-reported. Logistic regression models, adjusted for age, sex, BMI, Townsend Deprivation Index and other work factors, were used to investigate the relationships between work characteristics and osteoarthritis outcomes.

**Results:** This study included 285,947 participants (mean age 52.7 years; males 48.0%). Shift work and night shifts were associated with knee osteoarthritis (fully adjusted OR: 1.12 [95% CI:1.07-1.17] and 1.12 [1.04-1.20], respectively), and self-reported osteoarthritis but there was little evidence of an association with hip osteoarthritis (1.01 [0.95-1.08] and 1.03 [0.93-1.14]). Heavy manual work and prolonged non-sedentary work were associated with increased risk of all osteoarthritis outcomes.

**Conclusions:** Shift work showed independent associations with knee osteoarthritis and self-reported osteoarthritis but not hip osteoarthritis, suggesting circadian rhythm dysfunction may play a role in knee osteoarthritis pathogenesis. Heavy manual work and prolonged non-sedentary work were associated with all outcomes, with stronger associations in knee osteoarthritis, possibly reflecting the knee’s higher susceptibility to biomechanical stress. Further research is needed to explore workplace interventions for reducing these risks.

## Introduction

Osteoarthritis ranks as the 7^th^ leading cause of years lived with disability worldwide, particularly in those aged over 70, with the knee and hip being the most affected joints (1, 2). The global disease burden is increasing due to an upsurge in prevalence, driven by an aging demographic and a growing incidence of obesity (3). The identification of risk factors has the potential to alleviate this burden by helping prevent disease onset and progression. Given that individuals in the UK spend an average of 1532 hours at work annually (4), the workplace environment emerges as an important area for uncovering such risk factors. Notably, shift work-induced circadian disruption has received growing interest, with over 20% of European workers engaged in shift work in 2015 (5).

Circadian (24-hourly) rhythm has been shown to play a role in maintaining joint tissue homeostasis (6). Circadian rhythm is generated by a network of clock genes and proteins that rhythmically regulate their own expression and that of other tissue-specific target genes. This regulation occurs through a transcription-translation feedback loop (7). Notably, genes involved in extracellular matrix turnover and chondrocyte differentiation demonstrate circadian rhythmicity, with clock genes implicated in this regulation (8). In human osteoarthritis models, disruptions in clock gene function have been linked to increased expression of cartilage catabolic genes (9, 10), while environmental disturbances to circadian rhythms have been shown to induce osteoarthritis-like pathological alterations in the mouse knee joint (11).

Shift work is a known disruptor of circadian rhythms, by causing a misalignment in the sleep-wake cycle, raising the possibility it might influence osteoarthritis risk (12). Retrospective cohort studies of retired Chinese workers have revealed a positive association between shift work and risk of osteoarthritis, compared with daytime workers (13, 14). However, the generalisability of these findings to other populations may be limited. Of note, in the UK, both men and women engaging in night shift work are more likely to perform physically demanding tasks (15), which are a known risk factor for large joint osteoarthritis (16–18). Therefore, when exploring the association between shift work and osteoarthritis, it is important to consider the influence of work physicality as a possible confounding factor.

This study aimed to investigate the relationships between work characteristics and the prevalence of osteoarthritis within the UK Biobank (UKB) study. Specifically, our objectives were to explore the independent associations between shift work and the prevalence of hospital-diagnosed knee and hip osteoarthritis, as well as self-reported osteoarthritis, whilst considering the influence of work physicality. In addition, we sought to identify any sex differences in these relationships. By addressing these questions, our study aimed to offer insights into the associations between these work characteristics and joint health, with the goal of informing the development of future interventions and policies concerning the design of employment patterns, to alleviate the burden of osteoarthritis

## Materials and Methods

### Population

UKB is a longitudinal, population-based cohort study, that commenced in 2006, recruiting over 500,000 individuals aged 40-69 from across the UK. Participants underwent extensive physical and lifestyle assessment, including questionnaires on socio-demographic and environmental factors. Consent was given for linkage to electronic healthcare records, which enabled participant follow-up (19). UKB obtained ethical approval from the National Information Governance Board for Health and Social Care and Northwest Multi-centre Research Ethics Committee (11/NW/0382). All participants provided informed consent for the collection and use of their data.

### Self-reported Outcomes

Self-reported diagnoses of non-site-specific osteoarthritis were obtained through a questionnaire administered at baseline and during three follow-up periods. Participants were asked: *“Have you ever been told by a doctor that you have had osteoarthritis affecting one or more joints (e.g. hip, knee, shoulder)?”.* Participants could respond with “Yes”, “No”, “Do not know,” or “Prefer not to answer”. A binary variable was created by coding participants as “1” if they answered yes at any timepoint and as “0” if they did not answer yes at any point. Those who did not know or preferred not to answer were excluded from analyses.

### Diagnosed Outcomes

Hospital diagnosed knee osteoarthritis and hip osteoarthritis cases were identified through linkage to hospital episode statistics (HES), which use the International Classification of Diseases (ICD) 9^th^ and 10^th^ revision codes. Specifically, cases were identified using codes adopted from Zengini et al. (20) (Supplementary Table 1). The linkage of all UKB participants was conducted both prospectively and retrospectively at baseline. Records were available from 1st April 1997, and the data was downloaded from the UKB Showcase in August 2023, encompassing information until the end of October 2022.

### Exposures

Participants were asked about their work patterns at baseline and three follow-up periods, through a self-reported touchscreen questionnaire. This included “*Does your work involve shift work?*”, defined as a work schedule outside the routine daytime working hours of 09:00 to 17:00. Participants responded according to a four-point Likert scale: “Never/Rarely”, “Sometimes”, “Usually” or “Always”. Those who answered sometimes, usually or always were subsequently asked “*Does your work involve night shifts?*”, defined as a work schedule that involves working during the regular sleeping hours, for example from 00:00 to 06:00. Participants could answer according to the same four responses. For work physicality, participants were asked, “*Does your work involve heavy manual or physical work?*” and “*Does your work involve walking or standing for most of the time?*”. Heavy manual work was defined as employment involving the use of heavy objects and tools. Prolonged non-sedentary work was defined as work that requires prolonged periods of walking or standing. Again, responses followed the same four-category scale.

Categorical variables for the work exposures were created and coded: (1) never/rarely, (2) sometimes, (3) usually or (4) always. From this, a binary exposure variable was generated: (0) never/rarely and sometimes or (1) usually and always. Questionnaire data spanning the four timepoints was examined but considering most respondents only answered once, this information was collapsed into one set of variables. If a participant answered more than once, their highest response was selected. Those who did not know or preferred not to answer were excluded from analyses.

### Statistical Analysis

Descriptive statistics summarised the baseline population characteristics, presented as means, standard deviations (SDs) and ranges for continuous variables and as frequencies for categorical and binary variables. Logistic regression models examined the associations between each work exposure (shift work, night shifts, heavy manual work and prolonged non-sedentary work) and each osteoarthritis outcome (knee, hip and self-reported). Results are shown as odds ratios (OR) with 95% confidence intervals (CIs). Directed acyclic graphs guided the a priori selection of covariates for adjustment, specifically age, sex, body mass index (BMI) and Townsend Deprivation Index (TDI). The TDI is a postcode-based measure of socioeconomic deprivation, with a higher measure indicating increased socioeconomic deprivation (21). Models of shift work exposures were adjusted accordingly: model (1) unadjusted, (2) age and sex, (3) age, sex, BMI and TDI, (4) age, sex, BMI, TDI and both work physicality variables. Models using work physicality exposures were similarly adjusted except for model 4, which was adjusted for age, sex, BMI, TDI, shift work and the other work physicality variable. Age, BMI and TDI were incorporated into the models as continuous variables. Shift work and night shifts were not mutually adjusted due to direct classification overlap. The effect of exposure frequency on risk of osteoarthritis was explored using categorial work exposure variables, which used the “never” group as the comparator. Combined sex analyses were supplemented by sex-stratified analyses. Sex interactions terms were also examined. All statistical analyses were conducted using STATA version 18 (StataCorp, College Station, TX, USA).

## Results

### Baseline characteristics

In total, 285,947 participants (mean age 52.7, SD: 7.1 years, range: 38-71 years), were included in the analysis (Table 1). There were 137,154 (48.0%) males and 148,793 (52.0%) females. Knee osteoarthritis and hip osteoarthritis were diagnosed in 18,578 (6.5%) and 10,698 (3.7%) individuals, respectively. Non-site-specific osteoarthritis was self-reported in 16,407 (5.7%) of individuals. Knee osteoarthritis was more common among males with a prevalence of 9,623 (7.0%) compared to females (8,955 [6.0%]). Hip osteoarthritis and self-reported osteoarthritis were more common among females with a prevalence of 5,895 (4.0%) and 10,018 (6.7%). In males, 4,803 (3.5%) had hip osteoarthritis and 6,389 (4.7%) had self-reported osteoarthritis. Shift work, nights shifts and heavy manual work were more commonly reported among males, whereas prolonged non-sedentary work had a similar prevalence among males and females (Table1).

**Table 1.**
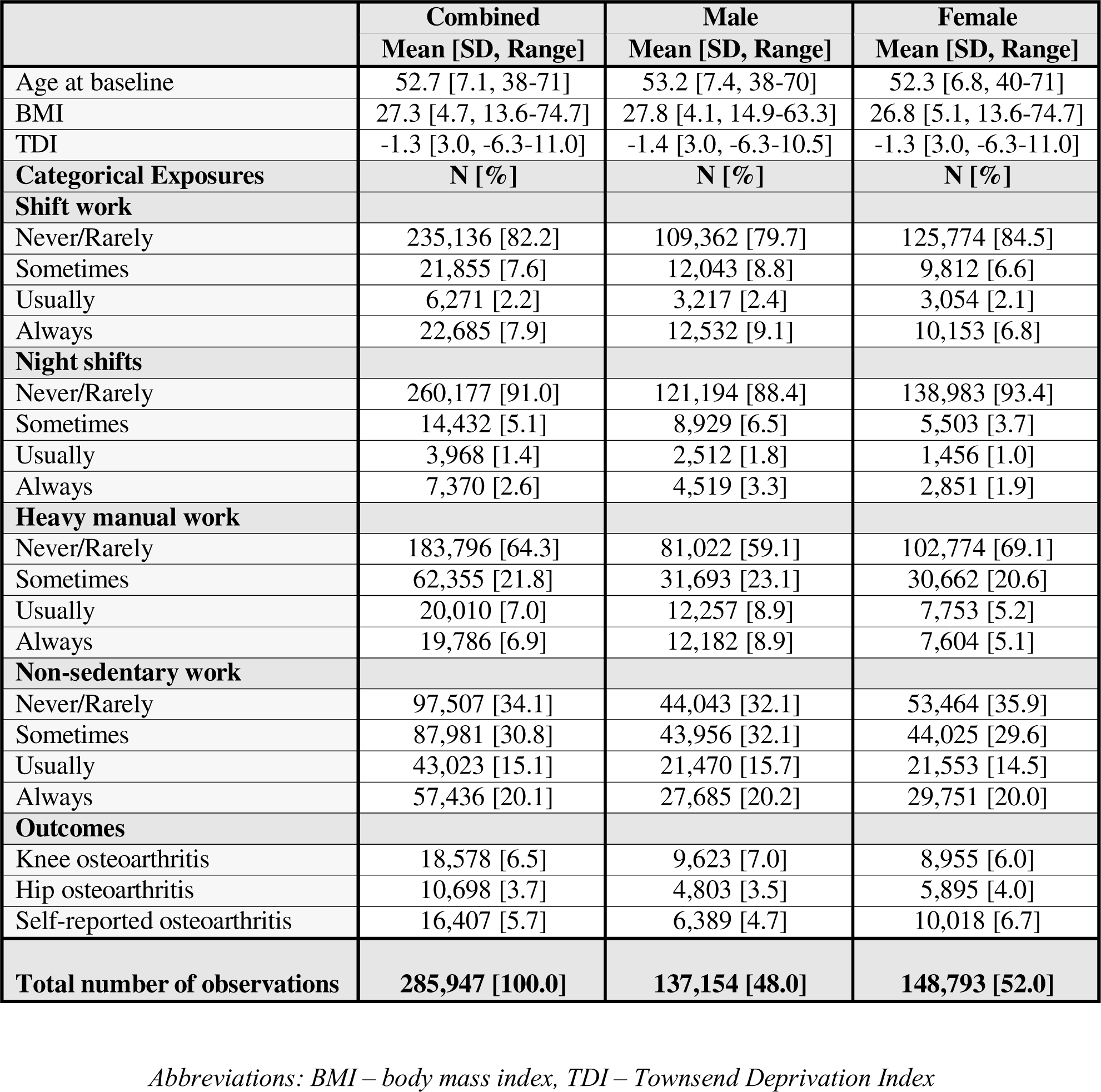
Descriptive statistics of study population.

|  | **Combined** | **Male** | **Female** |
| --- | --- | --- | --- |
|  | **Mean [SD, Range]** | **Mean [SD, Range]** | **Mean [SD, Range]** |
| Age at baseline | 52.7 [7.1, 38-71] | 53.2 [7.4, 38-70] | 52.3 [6.8, 40-71] |
| BMI | 27.3 [4.7, 13.6-74.7] | 27.8 [4.1, 14.9-63.3] | 26.8 [5.1, 13.6-74.7] |
| TDI | -1.3 [3.0, -6.3-11.0] | -1.4 [3.0, -6.3-10.5] | -1.3 [3.0, -6.3-11.0] |
| **Categorical Exposures** | **N [%]** | **N [%]** | **N [%]** |
| **Shift work** |  |  |  |
| Never/Rarely | 235,136 [82.2] | 109,362 [79.7] | 125,774 [84.5] |
| Sometimes | 21,855 [7.6] | 12,043 [8.8] | 9,812 [6.6] |
| Usually | 6,271 [2.2] | 3,217 [2.4] | 3,054 [2.1] |
| Always | 22,685 [7.9] | 12,532 [9.1] | 10,153 [6.8] |
| **Night shifts** |  |  |  |
| Never/Rarely | 260,177 [91.0] | 121,194 [88.4] | 138,983 [93.4] |
| Sometimes | 14,432 [5.1] | 8,929 [6.5] | 5,503 [3.7] |
| Usually | 3,968 [1.4] | 2,512 [1.8] | 1,456 [1.0] |
| Always | 7,370 [2.6] | 4,519 [3.3] | 2,851 [1.9] |
| **Heavy manual work** |  |  |  |
| Never/Rarely | 183,796 [64.3] | 81,022 [59.1] | 102,774 [69.1] |
| Sometimes | 62,355 [21.8] | 31,693 [23.1] | 30,662 [20.6] |
| Usually | 20,010 [7.0] | 12,257 [8.9] | 7,753 [5.2] |
| Always | 19,786 [6.9] | 12,182 [8.9] | 7,604 [5.1] |
| **Non-sedentary work** |  |  |  |
| Never/Rarely | 97,507 [34.1] | 44,043 [32.1] | 53,464 [35.9] |
| Sometimes | 87,981 [30.8] | 43,956 [32.1] | 44,025 [29.6] |
| Usually | 43,023 [15.1] | 21,470 [15.7] | 21,553 [14.5] |
| Always | 57,436 [20.1] | 27,685 [20.2] | 29,751 [20.0] |
| **Outcomes** |  |  |  |
| Knee osteoarthritis | 18,578 [6.5] | 9,623 [7.0] | 8,955 [6.0] |
| Hip osteoarthritis | 10,698 [3.7] | 4,803 [3.5] | 5,895 [4.0] |
| Self-reported osteoarthritis | 16,407 [5.7] | 6,389 [4.7] | 10,018 [6.7] |
| **Total number of observations** | **285,947 [100.0]** | **137,154 [48.0]** | **148,793 [52.0]** |

*Abbreviations: BMI – body mass index, TDI – Townsend Deprivation Index*

### Knee Osteoarthritis

Combined sex analyses, unadjusted for work physicality, demonstrated consistent associations between binary shift work and knee osteoarthritis (model 1: odds ratio (OR) 1.31 [95% CI: 1.25-1.37], model 2: 1.42 [1.36-1.49], model 3: 1.29 [1.23-1.35]). A similar trend was seen for night shifts (Table 2, Supplementary Figure 1). In sex stratified analyses, the effect estimates were comparable between males and females and the inclusion of a sex interaction term in the models did not provide strong evidence of an interaction (p<0.1), except in the unadjusted shift work model (Table 2, Supplementary Figure 1). Work physicality exposures exhibited stronger associations than the shift work exposures, with the point estimates for heavy manual work (model 1: OR 1.55 [95% CI: 1.49-1.61], model 2: 1.59 [1.53-1.65], model 3: 1.58 [1.52-1.64]) slightly higher than prolonged non-sedentary work (model 1: 1.48 [1.44-1.53], model 2: 1.46 [1.41-1.50], model 3: 1.48 [1.43-1.52]). For both work physicality exposures, sex interactions were seen in all models and sex stratification indicated that heavy manual work and prolonged non-sedentary work were more strongly associated with knee osteoarthritis in males compared to females (Table 2, Supplementary Figure 1).

**Table 2.** Logistic regression results showing the associations between binary work exposures and knee osteoarthritis in combined and sex-stratified analyses.

| Knee Osteoarthritis | Model 1 |  | Model 2 |  | Model 3 |  | Model 4 |  |
| --- | --- | --- | --- | --- | --- | --- | --- | --- |
|  | OR [95% CI] | P | OR [95% CI] | P | OR [95% CI] | P | OR [95% CI] | P |
| **Combined** |  |  |  |  |  |  |  |  |
| **Shift work** | 1.31 [1.25-1.37] | $8.87 \times 10^{-32*}$ | 1.42 [1.36-1.49] | $6.30 \times 10^{-52}$ | 1.29 [1.23-1.35] | $3.73 \times 10^{-27}$ | 1.12 [1.07-1.17] | $3.87 \times 10^{-6*}$ |
| **Night shifts** | 1.32 [1.23-1.42] | $1.26 \times 10^{-15}$ | 1.46 [1.36-1.57] | $6.61 \times 10^{-27}$ | 1.30 [1.21-1.39] | $3.39 \times 10^{-13}$ | 1.12 [1.04-1.20] | $1.75 \times 10^{-3*}$ |
| **Heavy manual work** | 1.55 [1.49-1.61] | $2.00 \times 10^{-113*}$ | 1.59 [1.53-1.65] | $3.00 \times 10^{-122*}$ | 1.58 [1.52-1.64] | $4.00 \times 10^{-114*}$ | 1.30 [1.24-1.36] | $5.76 \times 10^{-30*}$ |
| **Non-sedentary work** | 1.48 [1.44-1.53] | $2.00 \times 10^{-146*}$ | 1.46 [1.41-1.50] | $7.00 \times 10^{-131*}$ | 1.48 [1.43-1.52] | $3.00 \times 10^{-135*}$ | 1.32 [1.28-1.37] | $5.30 \times 10^{-54*}$ |
| **Males** |  |  |  |  |  |  |  |  |
| **Shift work** | 1.24 [1.16-1.31] | $9.11 \times 10^{-12}$ | 1.39 [1.31-1.48] | $6.79 \times 10^{-26}$ | 1.32 [1.24-1.40] | $1.09 \times 10^{-17}$ | 1.14 [1.07-1.21] | $8.34 \times 10^{-5}$ |
| **Night shifts** | 1.24 [1.14-1.35] | $1.14 \times 10^{-6}$ | 1.43 [1.31-1.56] | $1.41 \times 10^{-15}$ | 1.34 [1.23-1.46] | $1.14 \times 10^{-10}$ | 1.15 [1.05-1.26] | $2.14 \times 10^{-3}$ |
| **Heavy manual work** | 1.65 [1.57-1.73] | $4.70 \times 10^{-93}$ | 1.73 [1.65-1.81] | $2.00 \times 10^{-108}$ | 1.76 [1.68-1.85] | $3.00 \times 10^{-112}$ | 1.38 [1.30-1.46] | $6.18 \times 10^{-28}$ |
| **Non-sedentary work** | 1.66 [1.59-1.73] | $6.00 \times 10^{-126}$ | 1.64 [1.58-1.71] | $3.00 \times 10^{-119}$ | 1.69 [1.62-1.76] | $3.00 \times 10^{-127}$ | 1.45 [1.38-1.52] | $1.10 \times 10^{-46}$ |
| **Females** |  |  |  |  |  |  |  |  |
| **Shift work** | 1.38 [1.29-1.47] | $1.65 \times 10^{-20}$ | 1.45 [1.36-1.55] | $7.40 \times 10^{-27}$ | 1.26 [1.18-1.36] | $4.59 \times 10^{-11}$ | 1.14 [1.06-1.22] | $6.78 \times 10^{-4}$ |
| **Night shifts** | 1.39 [1.24-1.55] | $9.06 \times 10^{-9}$ | 1.49 [1.33-1.67] | $3.30 \times 10^{-12}$ | 1.23 [1.10-1.38] | $4.09 \times 10^{-4}$ | 1.09 [0.97-1.23] | 0.14 |
| **Heavy manual work** | 1.32 [1.24-1.41] | $1.48 \times 10^{-17}$ | 1.38 [1.29-1.47] | $3.82 \times 10^{-22}$ | 1.31 [1.23-1.40] | $6.39 \times 10^{-16}$ | 1.12 [1.04-1.21] | $1.97 \times 10^{-3}$ |
| **Non-sedentary work** | 1.31 [1.25-1.37] | $1.26 \times 10^{-33}$ | 1.28 [1.22-1.33] | $1.32 \times 10^{-27}$ | 1.28 [1.23-1.34] | $1.62 \times 10^{-27}$ | 1.22 [1.16-1.28] | $1.76 \times 10^{-14}$ |

Adjusted accordingly: Model 1 – unadjusted, Model 2 – age and sex, Model 3 – age, sex, BMI and TDI, Model 4 – age, sex, BMI, TDI and other employment factors.
\* Denotes a sex interaction term with P-value <0.1.

Mutual adjustment (model 4) attenuated the associations between binary work characteristics and knee osteoarthritis. In combined sex analyses shift work and night shifts continued to be associated with knee osteoarthritis (shift work: OR 1.12 [95% CI: 1.07-1.17], night shifts: 1.12 [1.04-1.20]). Unlike models 1-3, sex interactions were noted in the fully adjusted models for shift work and night shifts (Table 2). After sex stratification, night shifts remained associated with knee osteoarthritis in males (1.15 [1.05-1.26]) but weakly in females (1.09 [0.97-1.23]) Similarly, in combined sex analyses, work physicality remained strongly associated with knee osteoarthritis in the mutually adjusted model (heavy manual work: 1.30 [1.24-1.36], prolonged non-sedentary work: 1.32 [1.28-1.37]). Sex interactions persisted, with once again stronger associations seen between work physicality and knee osteoarthritis in males than in females (Table 2, Supplementary Figure 1).

When assessing work type as a categorical variable (1–4) in fully adjusted models, there was no indication of a progressive positive association between both more frequent shift work and nights shifts and knee osteoarthritis, with similar results seen in the sex stratified analyses (Figure 1, Supplementary Table 2). A distinct progressive association emerged between frequency of heavy manual work and knee osteoarthritis (sometimes: 1.17 [1.12-1.22], usually: 1.34 [1.26-1.42], always: 1.45 [1.36-1.55]), which was also the case for prolonged non-sedentary work (sometimes: 1.17 [1.13-1.22], usually: 1.33 [1.26-1.40], always: 1.38 [1.31-1.46]). The progressive trend was reflected in results for males and females, but effect sizes were larger in males (Figure 1, Supplementary Table 2).

**Figure 1.**
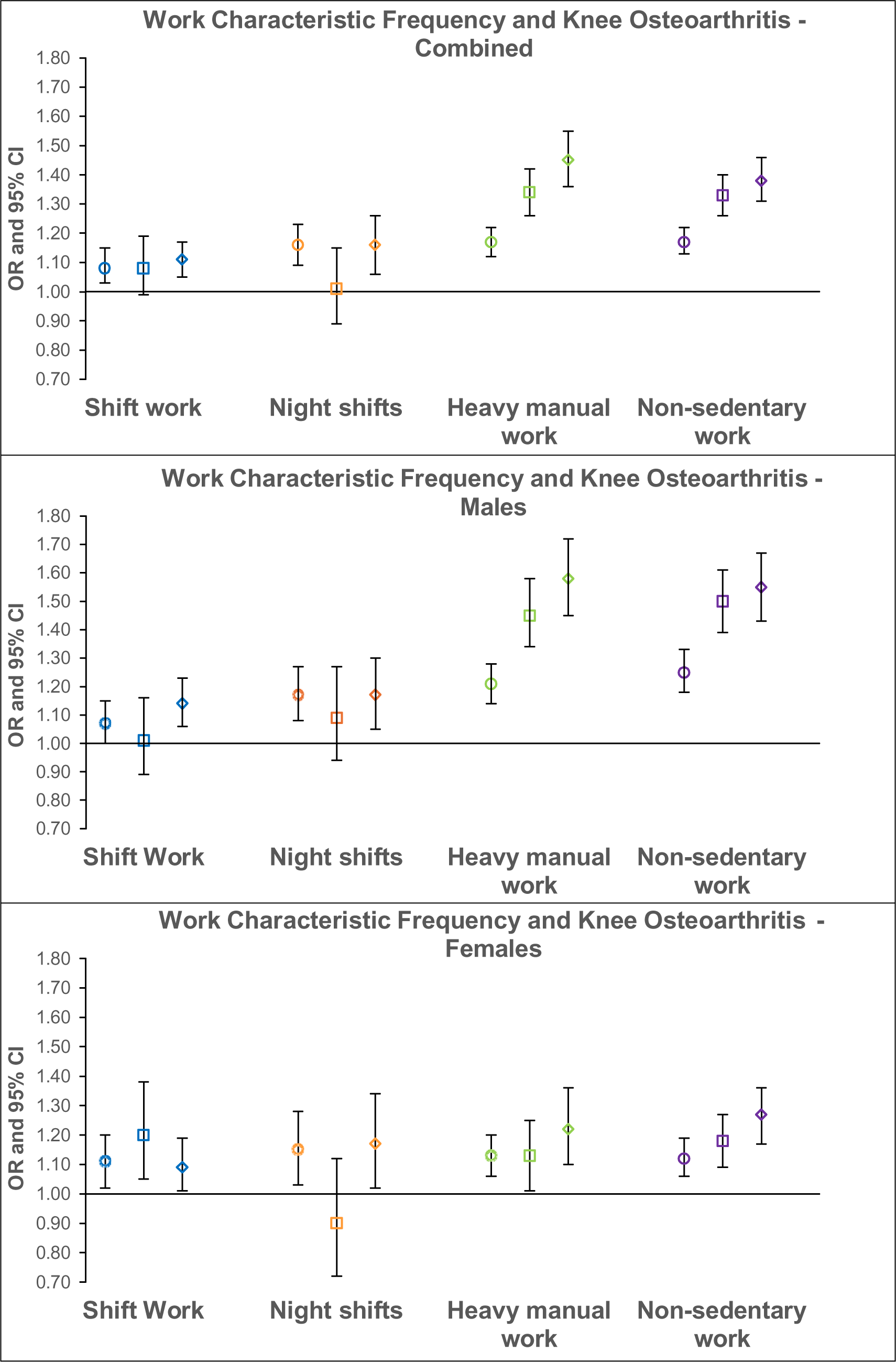
Logistic regression results for the associations between categorical work characteristic frequency and knee osteoarthritis in fully-adjusted combined and sex-stratified analyses. Odds ratios with the comparator group “never” are displayed with 95% confidence intervals. Different shapes represent the various employment intensities - circle: “sometimes”, square: “usually” and diamond: “always”.

### Hip Osteoarthritis

Combined sex analyses, unadjusted for work physicality, demonstrated little evidence of associations between binary shift work and hip osteoarthritis (model 1: OR 0.99 [95% CI: 0.93-1.05], model 2: 1.12 [1.05-1.20], model 3: 1.08 [1.01-1.15]). A similar lack of association was seen in analyses of night shifts. No sex interactions were seen for either exposure across models 1-3 and sex-stratified analyses indicated little differences between males and females (Table 3, Supplementary Figure 2). More robust associations were seen between work physicality and hip osteoarthritis, although weaker than those for knee osteoarthritis (Table 3, Supplementary Figure 2). Sex interactions were only evident for prolonged non-sedentary work. Sex-stratified analyses saw s t r on ge r associations between prolonged non-sedentary work and hip osteoarthritis in males (model 1: 1.26 [1.19-1.34], model 2: 1.24 [1.17-1.31], model 3: 1.26 [1.18-1.33]) than females (model 1: 1.14 [1.08-1.20], model 2: 1.10 [1.04-1.16] model 3: 1.10 [1.04-1.16]) (Table 3, Supplementary Figure 2).

**Table 3.** Logistic regression results showing the associations between binary work exposures and hip osteoarthritis in combined and sex-stratified analyses.

| Hip Osteoarthritis | Model 1 |  | Model 2 |  | Model 3 |  | Model 4 |  |
| --- | --- | --- | --- | --- | --- | --- | --- | --- |
|  | OR [95% CI] | P | OR [95% CI] | P | OR [95% CI] | P | OR [95% CI] | P |
| **Combined** |  |  |  |  |  |  |  |  |
| **Shift work** | 0.99 [0.93-1.05] | 0.74 | 1.12 [1.05-1.20] | $5.60 \times 10^{-4}$ | 1.08 [1.01-1.15] | 0.03 | 1.01 [0.95-1.08] | 0.74 |
| **Night shifts** | 0.97 [0.88-1.07] | 0.54 | 1.16 [1.04-1.28] | $5.24 \times 10^{-3}$ | 1.10 [0.99-1.21] | 0.08 | 1.03 [0.93-1.14] | 0.59* |
| **Heavy manual work** | 1.13 [1.07-1.19] | $1.00 \times 10^{-5}$ | 1.22 [1.15-1.29] | $1.55 \times 10^{-12}$ | 1.22 [1.15-1.28] | $2.87 \times 10^{-12}$ | 1.13 [1.06-1.20] | $1.32 \times 10^{-4}$ |
| **Non-sedentary work** | 1.19 [1.14-1.24] | $5.10 \times 10^{-18}$* | 1.16 [1.11-1.21] | $3.53 \times 10^{-13}$* | 1.17 [1.12-1.21] | $5.17 \times 10^{-14}$* | 1.12 [1.07-1.17] | $1.39 \times 10^{-6}$* |
| **Males** |  |  |  |  |  |  |  |  |
| **Shift work** | 0.96 [0.88-1.05] | 0.39 | 1.12 [1.02-1.23] | 0.01 | 1.09 [0.99-1.19] | 0.08 | 1.02 [0.93-1.12] | 0.67 |
| **Night shifts** | 0.99 [0.86-1.12] | 0.83 | 1.19 [1.04-1.36] | $9.45 \times 10^{-3}$ | 1.15 [1.01-1.31] | 0.04 | 1.08 [0.94-1.23] | 0.27 |
| **Heavy manual work** | 1.16 [1.08-1.25] | $3.96 \times 10^{-5}$ | 1.23 [1.14-1.32] | $3.40 \times 10^{-8}$ | 1.24 [1.15-1.33] | $7.12 \times 10^{-9}$ | 1.10 [1.01-1.20] | 0.03 |
| **Non-sedentary work** | 1.26 [1.19-1.34] | $5.51 \times 10^{-15}$ | 1.24 [1.17-1.31] | $1.34 \times 10^{-12}$ | 1.26 [1.18-1.33] | $5.04 \times 10^{-14}$ | 1.21 [1.13-1.29] | $1.18 \times 10^{-7}$ |
| **Females** |  |  |  |  |  |  |  |  |
| **Shift work** | 1.04 [0.95-1.13] | 0.43 | 1.11 [1.01-1.21] | 0.03 | 1.06 [0.96-1.16] | 0.25 | 1.00 [0.91-1.10] | 0.95 |
| **Night shifts** | 0.99 [0.85-1.16] | 0.90 | 1.08 [0.93-1.27] | 0.31 | 1.01 [0.86-1.19] | 0.88 | 0.95 [0.81-1.11] | 0.53 |
| **Heavy manual work** | 1.14 [1.05-1.24] | $1.53 \times 10^{-3}$ | 1.20 [1.11-1.30] | $1.38 \times 10^{-5}$ | 1.19 [1.09-1.29] | $5.53 \times 10^{-5}$ | 1.14 [1.04-1.25] | $4.75 \times 10^{-3}$ |
| **Non-sedentary work** | 1.14 [1.08-1.20] | $2.53 \times 10^{-6}$ | 1.10 [1.04-1.16] | $8.64 \times 10^{-4}$ | 1.10 [1.04-1.16] | $7.82 \times 10^{-4}$ | 1.06 [1.00-1.13] | 0.06 |

Adjusted accordingly: Model 1 – unadjusted, Model 2 – age and sex, Model 3 – age, sex, BMI and TDI, Model 4 – age, sex, BMI, TDI and other employment factors.
\* Denotes a sex interaction term with P-value <0.1.

Mutual adjustment (model 4) further attenuated the associations between binary work characteristics and hip osteoarthritis towards the null (shift work: OR 1.01 [95% CI: 0.95-1.08], night shifts: 1.03 [0.93-1.14]). In sex-stratified analysis, a similar lack of association between hip osteoarthritis and shift work was seen in males and females (Table 3, Supplementary Figure 2). In mutually adjusted combined sex analyses, associations between work physicality and hip osteoarthritis remained (heavy manual work: 1.13 [1.06-1.20], prolonged non-sedentary work: 1.12 [1.07-1.17]). Heavy manual work was similarly associated with hip osteoarthritis in females (1.14 [1.04-1.25]) and males (1.10 [1.01-1.20]). Sex interactions persisted for prolonged non-sedentary work, with stronger associations in males (1.21 [1.13-1.29]) compared to females (1.06 [1.00-1.13]) (Table 3, Supplementary Figure 2).

When assessing work type as a categorical variable (1–4) in fully adjusted models, as with knee osteoarthritis, there was no indication of a progressive positive association between both more frequent shift work and nights shifts and hip osteoarthritis. Similar results were seen in sex stratified analysis (Figure 2, Supplementary Table 3). A more progressive association was notable between heavy manual work frequency and hip osteoarthritis (sometimes: 1.03 [0.98-1.09], usually: 1.15 [1.06-1.25], always: 1.15 [1.05-1.26]) and prolonged non-sedentary work frequency and hip osteoarthritis (sometimes: 1.14 [1.09-1.20], usually: 1.19 [1.11-1.27], always: 1.18 [1.10-1.27]). Stronger associations between both heavy manual work and prolonged non-sedentary work and hip osteoarthritis were evident in males. Interestingly, the progressive positive association between work physical frequency and risk of hip osteoarthritis was absent in females (Figure 2, Supplementary Table 3).

**Figure 2.**
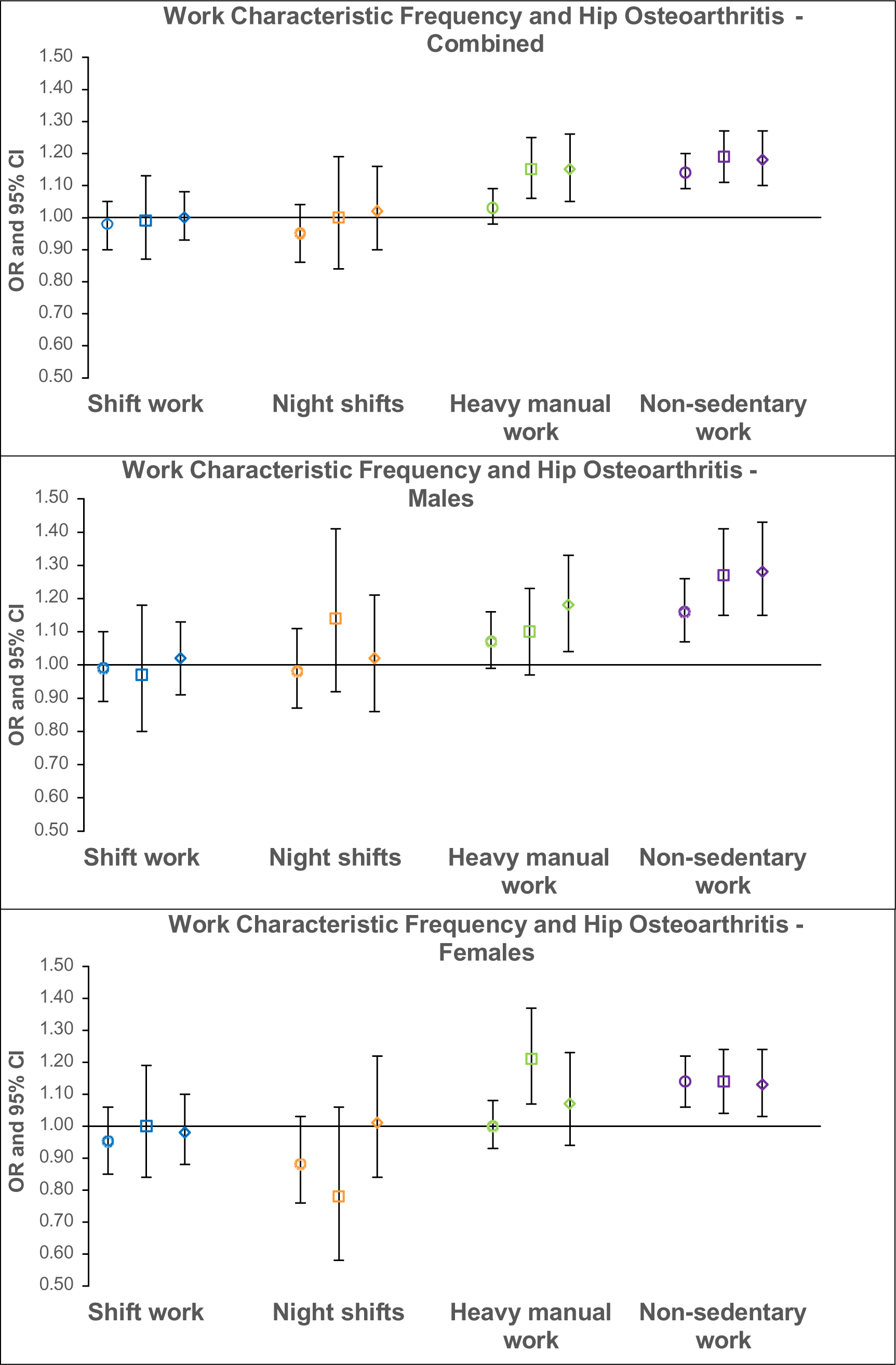
Logistic regression results for the associations between categorical work characteristic frequency and hip osteoarthritis in fully-adjusted combined and sex-stratified analyses. Odds ratios with the comparator group “never” are displayed with 95% confidence intervals. Different shapes represent the various employment intensities - circle: “sometimes”, square: “usually” and diamond: “always”.

### Self-reported Osteoarthritis

In combined sex analyses, unadjusted for work physicality, associations between binary shift work and self-reported osteoarthritis were present across all models (model 1: OR 1.15 [95% CI: 1.09-1.21], model 2: 1.32 [1.25-1.39], model 3: 1.23 [1.17-1.30]). Similar patterns emerged in night shift analyses. For both, stronger associations were seen in males and sex interactions were present in model 3, which incorporated adjustments for BMI and TDI alongside age and sex (Table 4, Supplementary Figure 3). In combined sex analyses, unadjusted for shift work, heavy manual work showed strong associations with self-reported osteoarthritis (model 1: OR 1.21 [95% CI: 1.16-1.26], model 2: 1.36 [1.30-1.42], model 3: 1.34 [1.28-1.40]). Results for prolonged non-sedentary work were similar. Like knee osteoarthritis, sex interactions were observed in all models for heavy manual work and prolonged non-sedentary work and in sex stratified analyses males exhibited higher odds of self-reported osteoarthritis (Table 4, Supplementary Figure 3).

**Table 4.** Logistic regression results showing the associations between binary work exposures and self-reported osteoarthritis in combined and sex-stratified analyses.

| Self-reported Osteoarthritis | Model 1 |  | Model 2 |  | Model 3 |  | Model 4 |  |
| --- | --- | --- | --- | --- | --- | --- | --- | --- |
|  | OR [95% CI] | P | OR [95% CI] | P | OR [95% CI] | P | OR [95% CI] | P |
| **Combined** |  |  |  |  |  |  |  |  |
| **Shift work** | 1.15 [1.09-1.21] | $6.83 \times 10^{-8}$ | 1.32 [1.25-1.39] | $1.16 \times 10^{-26}$ | 1.23 [1.17-1.30] | $1.51 \times 10^{-15*}$ | 1.13 [1.07-1.19] | $5.54 \times 10^{-6*}$ |
| **Night shifts** | 1.11 [1.03-1.20] | $6.20 \times 10^{-3}$ | 1.37 [1.27-1.48] | $3.56 \times 10^{-15}$ | 1.26 [1.16-1.36] | $1.22 \times 10^{-8*}$ | 1.15 [1.06-1.24] | $7.00 \times 10^{-4*}$ |
| **Heavy manual work** | 1.21 [1.16-1.26] | $1.21 \times 10^{-17*}$ | 1.36 [1.30-1.42] | $7.30 \times 10^{-43*}$ | 1.34 [1.28-1.40] | $2.95 \times 10^{-38*}$ | 1.20 [1.14-1.27] | $5.54 \times 10^{-13*}$ |
| **Non-sedentary work** | 1.26 [1.22-1.30] | $5.30 \times 10^{-46*}$ | 1.24 [1.20-1.28] | $4.96 \times 10^{-38*}$ | 1.24 [1.20-1.28] | $2.87 \times 10^{-37*}$ | 1.15 [1.10-1.19] | $1.27 \times 10^{-12*}$ |
| **Males** |  |  |  |  |  |  |  |  |
| **Shift work** | 1.19 [1.11-1.28] | $3.64 \times 10^{-6}$ | 1.38 [1.28-1.49] | $2.78 \times 10^{-17}$ | 1.31 [1.22-1.42] | $2.82 \times 10^{-12}$ | 1.20 [1.11-1.30] | $3.77 \times 10^{-6}$ |
| **Night shifts** | 1.16 [1.04-1.29] | $5.85 \times 10^{-3}$ | 1.39 [1.25-1.55] | $2.65 \times 10^{-9}$ | 1.31 [1.17-1.46] | $1.35 \times 10^{-6}$ | 1.19 [1.07-1.33] | $1.78 \times 10^{-3}$ |
| **Heavy manual work** | 1.35 [1.27-1.44] | $8.48 \times 10^{-23}$ | 1.43 [1.34-1.52] | $3.35 \times 10^{-30}$ | 1.42 [1.34-1.51] | $4.80 \times 10^{-29}$ | 1.21 [1.12-1.30] | $2.94 \times 10^{-7}$ |
| **Non-sedentary work** | 1.41 [1.34-1.49] | $1.00 \times 10^{-40}$ | 1.39 [1.32-1.46] | $1.57 \times 10^{-36}$ | 1.39 [1.32-1.46] | $4.00 \times 10^{-36}$ | 1.26 [1.19-1.34] | $7.61 \times 10^{-14}$ |
| **Females** |  |  |  |  |  |  |  |  |
| **Shift work** | 1.17 [1.09-1.25] | $4.84 \times 10^{-6}$ | 1.25 [1.17-1.34] | $1.15 \times 10^{-10}$ | 1.16 [1.08-1.24] | $3.00 \times 10^{-5}$ | 1.08 [1.00-1.16] | 0.04 |
| **Night shifts** | 1.20 [1.07-1.34] | $1.35 \times 10^{-3}$ | 1.32 [1.18-1.48] | $1.64 \times 10^{-6}$ | 1.18 [1.06-1.33] | $4.03 \times 10^{-3}$ | 1.09 [0.97-1.23] | 0.15 |
| **Heavy manual work** | 1.22 [1.15-1.30] | $3.33 \times 10^{-10}$ | 1.29 [1.21-1.37] | $4.64 \times 10^{-15}$ | 1.26 [1.18-1.34] | $3.19 \times 10^{-12}$ | 1.17 [1.09-1.26] | $1.42 \times 10^{-5}$ |
| **Non-sedentary work** | 1.18 [1.14-1.23] | $2.16 \times 10^{-15}$ | 1.15 [1.10-1.19] | $3.68 \times 10^{-10}$ | 1.14 [1.10-1.19] | $7.26 \times 10^{-10}$ | 1.08 [1.03-1.14] | $1.07 \times 10^{-3}$ |

Adjusted accordingly: Model 1 – unadjusted, Model 2 – age and sex, Model 3 – age, sex, BMI and TDI, Model 4 – age, sex, BMI, TDI and other employment factors.
\* Denotes a sex interaction term with P-value <0.1.

Like the other osteoarthritis outcomes, mutual adjustment for the other work factors (model 4) weakened the associations between binary work type and self-reported osteoarthritis. In combined sex analyses, shift work and night shifts retained associations with self-reported osteoarthritis (shift work: OR 1.13 [95% CI: 1.07-1.19], night shifts: 1.15 [1.06-1.24]). Sex interactions persisted for both (Table 4). Shift work and night shifts remained more strongly associated with self-reported osteoarthritis in males (shift work: 1.20 [1.11-1.30], night shifts: 1.19 [1.07-1.33]) than in females (shift work: 1.08 [1.00-1.16], night shifts: 1.09 [0.97-1.23]). Work physicality maintained its associations with self-reported osteoarthritis (heavy manual work: 1.20 [1.14-1.27], prolonged non-sedentary work: 1.15 [1.10-1.19]). Sex interactions remained, and again stronger associations were seen between work physicality and self-reported osteoarthritis in males than females (Table 4, Supplementary Figure 3).

Overall, in fully adjusted categorical analyses of shift work and night shift frequency, there were no progressive association with self-reported osteoarthritis (Figure 3, Supplementary Table 3). Regarding heavy manual work frequency, progressive stronger associations with self-reported osteoarthritis were observed in fully adjusted models (sometimes: 1.16 [1.11-1.22], usually: 1.26 [1.18-1.35], always: 1.33 [1.24-1.44]). Prolonged non-sedentary work displayed a similar progressive trend. Unlike knee and hip osteoarthritis, increasing heavy manual work frequency, saw progressive associations with self-reported osteoarthritis in females. In contrast, for prolonged non-sedentary work, progressive associations were seen in males but not females (Figure 3, Supplementary Table 3).

**Figure 3.**
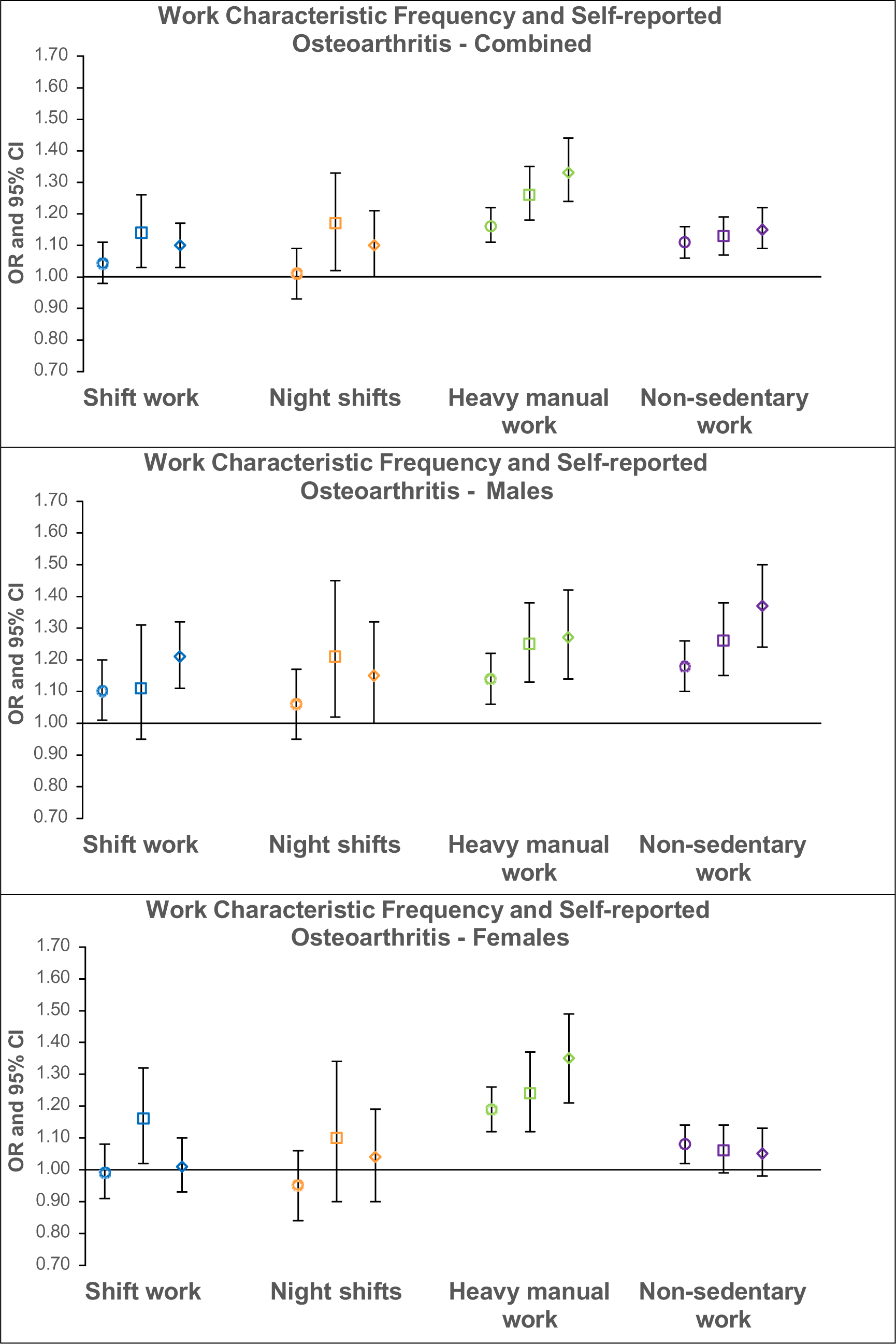
Logistic regression results for the associations between categorical work characteristic frequency and self-reported osteoarthritis in fully-adjusted combined and sex-stratified analyses. Odds ratios with the comparator group “never” are displayed with 95% confidence intervals. Different shapes represent the various employment intensities - circle: “sometimes”, square: “usually” and diamond: “always”.

## Discussion

As far as we are aware, this is the largest cross-sectional study to date exploring the relationships between work characteristics (i.e. shift work, night-shifts, heavy manual work and prolonged non-sedentary work) and osteoarthritis (knee, hip and self-reported). Overall, both shift work and night shifts were associated with an increased risk of knee osteoarthritis and self-reported osteoarthritis, while no such associations were observed for hip osteoarthritis. Importantly, these associations remained after adjustment for possible confounding factors including age, sex, BMI and deprivation, as well as for the physical nature of work. Greater associations were seen between work physicality and osteoarthritis compared to shift work and night shifts, which were stronger in the knee than the hip. Broadly, work characteristics showed larger association with osteoarthritis in males as compared with females. The increasing frequency of heavy manual work and prolonged non-sedentary work showed clear progressive trends with an increasing risk of osteoarthritis. Shift work and night shifts did not display a similar trend.

Previous epidemiological research has linked shift work to an increased risk of osteoarthritis, in smaller Chinese cohorts. One study saw a hazard ratio of 1.19 for knee osteoarthritis in shift workers compared to non-shift workers, after adjusting for work posture but also working years (13). Another study saw a comparable OR of 1.22 for lower-limb osteoarthritis in shift workers compared to day workers, after adjusting for work posture (14). In comparison, this study saw an OR of 1.12 for both shift work and night shifts with knee osteoarthritis, after adjusting for work physicality. The lack of a progressive association between increasing frequency of shift work or night shifts with risk of osteoarthritis could be attributed to the limited number of responses in the “*usually*” group, which underpowered these analyses. Nevertheless, progressive positive associations were present in physicality analyses, suggesting these associations are weaker for shift work. Another explanation is circadian adaptation among more permanent shift workers. However, research indicates that less than 3% show complete adjustment with only 25% experiencing a beneficial degree of adjustment. Therefore, it is unlikely that most workers undergo sufficient synchronisation to benefit their health (22).

Although causal associations cannot be inferred from this cross-sectional study, the findings are supported by animal studies, where circadian rhythm disruption, whether through environmental disturbance or genetic deletion of clock genes, contributed to osteoarthritis-like changes (11, 23). While underlying mechanisms remain unclear, shift work is an established cause of circadian misalignment, due to frequent alterations in sleep-wake and light-dark cycles and has been shown to desynchronise clock gene expression (24). Therefore, shift work may be viewed as a proxy for circadian rhythm disruption and these findings provide evidence that circadian rhythm dysfunction is an important risk factor for osteoarthritis. Interestingly, genetic knockout animal studies only saw degenerative cartilage changes in the knee joint and not at the hip, as was seen in this study (23). Since work physicality was adjusted for, this suggests that the knee may be more vulnerable to the effects of circadian rhythm disruption, independent of more extreme physical stressors. Indeed existing research has indicated disparities in the cellular and molecular pathophysiology of knee and hip osteoarthritis, including differences in inflammatory processes (25), but further research is required to elucidate any differences within the context of shift work and circadian rhythm disruption.

Both heavy manual work and prolonged non-sedentary work exposures demonstrated associations with all osteoarthritis outcomes. Occupational physical stressors and risk of osteoarthritis have been more extensively explored. For instance, a longitudinal cohort study found heavy manual work was associated with an increased risk of knee osteoarthritis, double that of sedentary work (26). In another cross-sectional study, workers conducting highly physical work exhibited a nearly two-fold higher prevalence of symptomatic osteoarthritis, compared to those with less physical work (27). Multiple reviews have also concluded that occupations characterised by heavy physical workloads, repetitive actions and prolonged walking and standing are associated with increased risk of knee and hip osteoarthritis (28–30).

Although heavy manual work and prolonged non-sedentary work showed associations with both knee and hip osteoarthritis, these were stronger at the knee. A different study also saw a greater association between job-related physical activity and knee osteoarthritis, with a hazard ratio of 1.39 compared with 1.22 for hip osteoarthritis (31). This may be explained by the increased susceptibility of the knee to biomechanical stress. During everyday movements the knee experiences higher peak compressive forces compared to the hip (32). Moreover, as the knee has more restricted movement and less stability, it is more inclined to injury, which can accelerate degenerative changes (25, 33).

Despite associations being present for both males and females, work physicality mostly displayed stronger associations with osteoarthritis in males. Similarly, a meta-analysis of 71 studies found greater odds of knee osteoarthritis in workers with physically demanding occupations, that were male (OR 1.61), compared to female (1.35) (28). Across all occupations, men demonstrate a stronger correlation between physical stressors, repetitive tasks, work demands and risk of injury (34). Moreover, in hip osteoarthritis analyses, the progressive associations between work frequency and risk of osteoarthritis were not observed in females and similar findings have been reported in other studies where positive associations were solely found in males (35). These differences could be attributed to the greater engagement in manual labour among the male demographic, reflected by the larger number of male-only studies (35), but more work is justified to fully understand the reasons behind this.

This study has identified robust associations between work-related exposures and large-joint osteoarthritis, which have important implications for occupational health policies. Further research is needed to establish causal relationships through randomised trials of workplace interventions. For example, therapies and interventions that tackle sleep hygiene, shift timing and light exposures have demonstrated favourable results in improving sleep among shift workers and may have a positive impact on joint health (36). Considering that certain occupational exposures are likely unavoidable, it may be more feasible to examine dose-response associations to identify thresholds of safe working and to introduce closer monitoring or screening of musculoskeletal health in these workers. Another area of interest is the potential chronotherapeutic administration of prospective disease-modifying therapies and it may be useful to explore differences in medication timing between day and night shift workers (37).

A notable strength of this study is its large sample size, enhancing the statistical power of the analyses. Mutual-adjustment allowed for the estimation of independent associations between shift work and work physicality with risk of osteoarthritis, to be drawn, addressing a gap in prior research. UKB contains extensive lifestyle data, enabling adjustments for age, sex, BMI and deprivation, which have been associated with osteoarthritis and employment. Previously, socioeconomic status has been largely unconsidered (13, 14), but was included in this study. A main limitation of this study is its cross-sectional nature, preventing the inference of causal associations in isolation. Having osteoarthritis could influence choices about employment, as these individuals would be more likely to avoid physically demanding or prolonged non-sedentary jobs, which could lower their odds. For shift work, the triangulation between this study and animal studies does add to the causal hypothesis that circadian rhythm dysfunction causes knee osteoarthritis. The population within UKB predominantly comprises Caucasian participants and those who tend to be more healthy and less deprived, restricting generalisability to other ethnicities. Additional research among more ethnically diverse cohorts would be beneficial. Another limitation is that this study was unable to account for employment duration (i.e. years worked) or regularity of shift patterns (e.g. fixed versus rotating shifts), which are likely to be important components of risk.

In conclusion, this large cross-sectional study saw independent associations between shift work and both knee and self-reported osteoarthritis, but not hip osteoarthritis. These findings support the role of circadian rhythm dysfunction as a contributing factor in knee osteoarthritis pathogenesis, which aligns with the results of previous animal studies. Although further replication of these findings is warranted given the duration of work exposures were not known. Work physicality demonstrated association with osteoarthritis at all sites, with the strongest associations seen in the knee, possibly reflecting its higher susceptibility to biomechanical stress. These findings highlight the importance of the workplace environment in risk factor management and could inform strategies to promote joint health in the workforce. Further research is warranted to explore the effects of workplace interventions in minimising the risk of osteoarthritis and to investigate the anatomical discrepancies that were observed.

## Data Availability

All data is available to registered users via UK Biobank showcase.

## Acknowledgements

This work has been conducted using the UK Biobank resource, access application 17295.

## Author contributions

Significant contributions were made by all authors towards the conception and design of this study, the acquisition of data, its analysis and interpretation. Each author helped draft the article before agreeing the final version of this manuscript. Asad Hashmi takes full responsibility for the integrity of the work.

