## Supplementary tables and figures for "Associations between work characteristics and large joint osteoarthritis: a cross-sectional study of 285,947 UK Biobank participants"

*Supplementary Table 1 - ICD 9 and 10 codes for osteoarthritis*

| | **ICD 10 codes** | | **ICD 9 codes** | |
|---|---|---|---|---|
| **Knee Osteoarthritis** | **M17** | Gonarthrosis [arthrosis of knee] | **71536** | Unspecific localised osteoarthrosis/allied disorder (lower leg) |
| | **M170** | Primary gonarthrosis, bilateral | | |
| | **M171** | Other primary gonarthrosis | | |
| | **M179** | Gonarthrosis, unspecified | **71516** | Localised, primary osteoarthrosis and allied disorders (lower leg) |
| | **M1906** | Primary arthrosis of other joints (lower leg) | | |
| | **M1996** | Arthrosis, unspecified (lower leg) | | |
| **Hip Osteoarthritis** | **M16** | Coxarthrosis [arthrosis of hip] | **71535** | Unspecified localised osteoarthrosis/allied disorder (pelvic region and thigh) |
| | **M160** | Primary coxarthrosis, bilateral | | |
| | **M161** | Other primary coxarthrosis | | |
| | **M169** | Coxarthrosis, unspecified | **71515** | Localised primary osteoarthrosis/allied disorder (pelvic region and thigh) |
| | **M1905** | Primary arthrosis of other joints (pelvic region and thigh) | | |
| | **M1995** | Arthrosis, unspecified (pelvic region and thigh) | | |

*Table adapted from Zengini et al. (20)*

*Supplementary Table 1 - Logistic regression results showing the associations between categorical work frequency and knee osteoarthritis in combined and sex-stratified analyses.*

| Knee Osteoarthritis | Shift Work | | | | Night Shifts | | | | Heavy Manual Work | | | | Non-sedentary Work | | | |
|---|---|---|---|---|---|---|---|---|---|---|---|---|---|---|---|---|
| | Unadjusted | | Fully Adjusted | | Unadjusted | | Fully Adjusted | | Unadjusted | | Fully Adjusted | | Unadjusted | | Fully Adjusted | |
| | OR [95%CI] | P | OR [95%CI] | P | OR [95%CI] | P | OR [95%CI] | P | OR [95%CI] | P | OR [95%CI] | P | OR [95%CI] | P | OR [95%CI] | P |
| **Combined** | | | | | | | | | | | | | | | | |
| **Sometimes** | 1.25 [1.19-1.32] | $4.08 \times 10^{-17}$ | 1.08 [1.03-1.15] | $3.89 \times 10^{-3}$ | 1.32 [1.24-1.40] | $9.52 \times 10^{-19}$ | 1.16 [1.09-1.23] | $8.28 \times 10^{-6}$ | 1.41 [1.36-1.46] | $1.50 \times 10^{-78}$ | 1.17 [1.12-1.22] | $3.77 \times 10^{-13}$ | 1.31 [1.26-1.36] | $1.10 \times 10^{-40}$ | 1.17 [1.13-1.22] | $4.46 \times 10^{-14}$ |
| **Usually** | 1.31 [1.20-1.44] | $9.49 \times 10^{-9}$ | 1.08 [0.99-1.19] | $9.76 \times 10^{-2}$ | 1.17 [1.04-1.32] | $9.67 \times 10^{-3}$ | 1.01 [0.89-1.15] | 0.85 | 1.65 [1.56-1.74] | $6.20 \times 10^{-77}$ | 1.34 [1.26-1.42] | $6.97 \times 10^{-20}$ | 1.58 [1.51-1.66] | $4.40 \times 10^{-85}$ | 1.33 [1.26-1.40] | $3.21 \times 10^{-26}$ |
| **Always** | 1.34 [1.28-1.41] | $1.26 \times 10^{-30}$ | 1.11 [1.05-1.17] | $2.87 \times 10^{-4}$ | 1.44 [1.32-1.56] | $3.86 \times 10^{-18}$ | 1.16 [1.06-1.26] | $7.55 \times 10^{-4}$ | 1.77 [1.68-1.86] | $8.00 \times 10^{-103}$ | 1.45 [1.36-1.55] | $1.19 \times 10^{-28}$ | 1.79 [1.72-1.87] | $2.00 \times 10^{-169}$ | 1.38 [1.31-1.46] | $3.88 \times 10^{-31}$ |
| **Males** | | | | | | | | | | | | | | | | |
| **Sometimes** | 1.23 [1.15-1.32] | $2.87 \times 10^{-9}$ | 1.07 [1.00-1.15] | 0.05 | 1.29 [1.19-1.39] | $1.63 \times 10^{-10}$ | 1.17 [1.08-1.27] | $1.02 \times 10^{-4}$ | 1.51 [1.44-1.59] | $6.20 \times 10^{-59}$ | 1.21 [1.14-1.28] | $3.35 \times 10^{-10}$ | 1.43 [1.35-1.51] | $7.41 \times 10^{-34}$ | 1.25 [1.18-1.33] | $4.78 \times 10^{-13}$ |
| **Usually** | 1.15 [1.01-1.31] | 0.04 | 1.01 [0.89-1.16] | 0.84 | 1.17 [1.01-1.35] | 0.04 | 1.09 [0.94-1.27] | 0.27 | 1.85 [1.73-1.97] | $5.10 \times 10^{-73}$ | 1.45 [1.34-1.58] | $6.30 \times 10^{-20}$ | 1.90 [1.78-2.02] | $1.00 \times 10^{-84}$ | 1.50 [1.39-1.61] | $4.48 \times 10^{-26}$ |
| **Always** | 1.29 [1.21-1.38] | $5.12 \times 10^{-14}$ | 1.14 [1.06-1.23] | $2.24 \times 10^{-4}$ | 1.32 [1.19-1.46] | $3.08 \times 10^{-7}$ | 1.17 [1.05-1.30] | $5.60 \times 10^{-3}$ | 1.92 [1.80-2.05] | $1.20 \times 10^{-84}$ | 1.58 [1.45-1.72] | $2.11 \times 10^{-25}$ | 2.10 [1.98-2.23] | $2.00 \times 10^{-133}$ | 1.55 [1.43-1.67] | $3.36 \times 10^{-27}$ |
| **Females** | | | | | | | | | | | | | | | | |
| **Sometimes** | 1.24 [1.14-1.34] | $2.23 \times 10^{-7}$ | 1.11 [1.02-1.20] | 0.02 | 1.30 [1.17-1.44] | $8.08 \times 10^{-7}$ | 1.15 [1.03-1.28] | $9.81 \times 10^{-3}$ | 1.29 [1.23-1.36] | $7.47 \times 10^{-23}$ | 1.13 [1.06-1.20] | $6.67 \times 10^{-5}$ | 1.21 [1.14-1.27] | $3.18 \times 10^{-11}$ | 1.12 [1.06-1.19] | $1.53 \times 10^{-4}$ |
| **Usually** | 1.49 [1.31-1.70] | $1.57 \times 10^{-9}$ | 1.20 [1.05-1.38] | $8.26 \times 10^{-3}$ | 1.09 [0.88-1.35] | 0.43 | 0.90 [0.72-1.12] | 0.34 | 1.32 [1.21-1.45] | $1.23 \times 10^{-9}$ | 1.13 [1.01-1.25] | 0.03 | 1.29 [1.21-1.38] | $3.60 \times 10^{-14}$ | 1.18 [1.09-1.27] | $1.70 \times 10^{-5}$ |
| **Always** | 1.37 [1.27-1.48] | $7.84 \times 10^{-16}$ | 1.09 [1.01-1.19] | 0.04 | 1.57 [1.38-1.79] | $1.46 \times 10^{-11}$ | 1.17 [1.02-1.34] | 0.03 | 1.50 [1.37-1.63] | $9.41 \times 10^{-20}$ | 1.22 [1.10-1.36] | $2.54 \times 10^{-4}$ | 1.53 [1.44-1.62] | $1.30 \times 10^{-46}$ | 1.27 [1.17-1.36] | $8.67 \times 10^{-10}$ |

*Odds ratios are in comparison to the "Never" group.*

*The fully adjusted model is adjusted for age, sex, BMI, TDI and other employment factors.*

***Supplementary Table 3 - Logistic regression results showing the associations between categorical work frequency and hip osteoarthritis in combined and sex-stratified analyses.***

| Hip Osteoarthritis | Shift Work | | | | Night Shifts | | | | Heavy Manual Work | | | | Non-sedentary Work | | | |
|---|---|---|---|---|---|---|---|---|---|---|---|---|---|---|---|---|
| | Unadjusted | | Fully Adjusted | | Unadjusted | | Fully Adjusted | | Unadjusted | | Fully Adjusted | | Unadjusted | | Fully Adjusted | |
| | OR [95%CI] | P | OR [95%CI] | P | OR [95%CI] | P | OR [95%CI] | P | OR [95%CI] | P | OR [95%CI] | P | OR [95%CI] | P | OR [95%CI] | P |
| **Combined** | | | | | | | | | | | | | | | | |
| **Sometimes** | 0.96 [0.90-1.04] | 0.34 | 0.98 [0.90-1.05] | 0.54 | 0.87 [0.79-0.96] | $3.66 \times 10^{-3}$ | 0.95 [0.86-1.04] | 0.26 | 1.11 [1.06-1.17] | $8.96 \times 10^{-6}$ | 1.03 [0.98-1.09] | 0.24 | 1.21 [1.15-1.27] | $3.59 \times 10^{-14}$ | 1.14 [1.09-1.20] | $2.66 \times 10^{-7}$ |
| **Usually** | 1.05 [0.92-1.19] | 0.49 | 0.99 [0.87-1.13] | 0.85 | 0.91 [0.76-1.08] | 0.26 | 1.00 [0.84-1.19] | 0.97 | 1.19 [1.11-1.29] | $1.75 \times 10^{-6}$ | 1.15 [1.06-1.25] | $1.24 \times 10^{-3}$ | 1.30 [1.22-1.38] | $8.90 \times 10^{-18}$ | 1.19 [1.11-1.27] | $2.37 \times 10^{-7}$ |
| **Always** | 0.97 [0.90-1.04] | 0.40 | 1.00 [0.93-1.08] | 0.96 | 0.99 [0.88-1.12] | 0.91 | 1.02 [0.90-1.16] | 0.70 | 1.13 [1.04-1.21] | $1.97 \times 10^{-3}$ | 1.15 [1.05-1.26] | $3.60 \times 10^{-3}$ | 1.32 [1.25-1.39] | $1.30 \times 10^{-23}$ | 1.18 [1.10-1.27] | $2.27 \times 10^{-6}$ |
| **Males** | | | | | | | | | | | | | | | | |
| **Sometimes** | 1.00 [0.90-1.11] | 0.98 | 0.99 [0.89-1.10] | 0.88 | 0.93 [0.82-1.04] | 0.21 | 0.98 [0.87-1.11] | 0.75 | 1.22 [1.14-1.31] | $1.49 \times 10^{-8}$ | 1.07 [0.99-1.16] | 0.09 | 1.27 [1.18-1.37] | $2.77 \times 10^{-10}$ | 1.16 [1.07-1.26] | $1.83 \times 10^{-4}$ |
| **Usually** | 0.97 [0.80-1.18] | 0.77 | 0.97 [0.80-1.18] | 0.77 | 1.03 [0.83-1.27] | 0.77 | 1.14 [0.92-1.41] | 0.24 | 1.23 [1.12-1.36] | $3.63 \times 10^{-5}$ | 1.10 [0.97-1.23] | 0.13 | 1.43 [1.31-1.57] | $1.13 \times 10^{-15}$ | 1.27 [1.15-1.41] | $3.68 \times 10^{-6}$ |
| **Always** | 0.96 [0.87-1.06] | 0.41 | 1.02 [0.91-1.13] | 0.77 | 0.95 [0.81-1.12] | 0.57 | 1.02 [0.86-1.21] | 0.81 | 1.24 [1.12-1.37] | $2.56 \times 10^{-5}$ | 1.18 [1.04-1.33] | 0.01 | 1.44 [1.32-1.56] | $5.87 \times 10^{-18}$ | 1.28 [1.15-1.43] | $6.63 \times 10^{-6}$ |
| **Females** | | | | | | | | | | | | | | | | |
| **Sometimes** | 0.95 [0.85-1.06] | 0.35 | 0.95 [0.85-1.06] | 0.36 | 0.83 [0.72-0.97] | 0.02 | 0.88 [0.76-1.03] | 0.11 | 1.05 [0.98-1.12] | 0.17 | 1.00 [0.93-1.08] | 0.97 | 1.18 [1.11-1.26] | $6.44 \times 10^{-7}$ | 1.14 [1.06-1.22] | $2.20 \times 10^{-4}$ |
| **Usually** | 1.13 [0.95-1.34] | 0.17 | 1.00 [0.84-1.19] | 0.98 | 0.77 [0.57-1.03] | 0.08 | 0.78 [0.58-1.06] | 0.11 | 1.24 [1.11-1.38] | $1.26 \times 10^{-4}$ | 1.21 [1.07-1.37] | $2.55 \times 10^{-3}$ | 1.21 [1.12-1.31] | $3.45 \times 10^{-6}$ | 1.14 [1.04-1.24] | $4.28 \times 10^{-3}$ |
| **Always** | 1.00 [0.91-1.11] | 0.94 | 0.98 [0.88-1.10] | 0.75 | 1.09 [0.91-1.31] | 0.33 | 1.01 [0.84-1.22] | 0.88 | 1.07 [0.95-1.20] | 0.28 | 1.07 [0.94-1.23] | 0.31 | 1.25 [1.16-1.34] | $2.51 \times 10^{-9}$ | 1.13 [1.03-1.24] | $9.88 \times 10^{-3}$ |

*Odds ratios are in comparison to the "Never" group.*

*The fully adjusted model is adjusted for age, sex, BMI, TDI and other employment factors.*

*Supplementary Table 4 - Logistic regression results showing the associations between categorical work frequency and self-reported osteoarthritis in combined and sex-stratified analyses.*

| Self-reported Osteoarthritis | Shift Work | | | | Night Shifts | | | | Heavy Manual Work | | | | Non-sedentary Work | | | |
|---|---|---|---|---|---|---|---|---|---|---|---|---|---|---|---|---|
| | Unadjusted | | Fully Adjusted | | Unadjusted | | Fully Adjusted | | Unadjusted | | Fully Adjusted | | Unadjusted | | Fully Adjusted | |
| | OR [95%CI] | P | OR [95%CI] | P | OR [95%CI] | P | OR [95%CI] | P | OR [95%CI] | P | OR [95%CI] | P | OR [95%CI] | P | OR [95%CI] | P |
| **Combined** | | | | | | | | | | | | | | | | |
| **Sometimes** | 1.06 [1.00-1.12] | 0.06 | 1.04 [0.98-1.11] | 0.16 | 0.95 [0.89-1.03] | 0.20 | 1.01 [0.93-1.09] | 0.86 | 1.24 [1.19-1.29] | $8.89 \times 10^{-29}$ | 1.16 [1.11-1.22] | $1.32 \times 10^{-11}$ | 1.20 [1.15-1.25] | $2.98 \times 10^{-18}$ | 1.11 [1.06-1.16] | $1.72 \times 10^{-6}$ |
| **Usually** | 1.25 [1.14-1.39] | $6.58 \times 10^{-6}$ | 1.14 [1.03-1.26] | 0.01 | 1.09 [0.96-1.24] | 0.20 | 1.17 [1.02-1.33] | 0.02 | 1.27 [1.20-1.35] | $4.44 \times 10^{-15}$ | 1.26 [1.18-1.35] | $4.43 \times 10^{-11}$ | 1.31 [1.24-1.37] | $4.99 \times 10^{-27}$ | 1.13 [1.07-1.19] | $1.97 \times 10^{-5}$ |
| **Always** | 1.12 [1.06-1.19] | $4.44 \times 10^{-5}$ | 1.10 [1.03-1.17] | $2.39 \times 10^{-3}$ | 1.12 [1.02-1.24] | 0.02 | 1.10 [1.00-1.21] | 0.06 | 1.29 [1.22-1.37] | $4.01 \times 10^{-17}$ | 1.33 [1.24-1.44] | $1.22 \times 10^{-14}$ | 1.44 [1.38-1.50] | $8.10 \times 10^{-60}$ | 1.15 [1.09-1.22] | $1.45 \times 10^{-6}$ |
| **Males** | | | | | | | | | | | | | | | | |
| **Sometimes** | 1.16 [1.07-1.26] | $6.21 \times 10^{-4}$ | 1.10 [1.01-1.20] | 0.03 | 1.06 [0.96-1.17] | 0.24 | 1.06 [0.95-1.17] | 0.28 | 1.36 [1.28-1.44] | $1.55 \times 10^{-23}$ | 1.14 [1.06-1.22] | $3.05 \times 10^{-4}$ | 1.33 [1.24-1.42] | $1.94 \times 10^{-16}$ | 1.18 [1.10-1.26] | $4.77 \times 10^{-6}$ |
| **Usually** | 1.17 [1.00-1.37] | 0.05 | 1.11 [0.95-1.31] | 0.19 | 1.18 [0.99-1.40] | 0.07 | 1.21 [1.02-1.45] | 0.03 | 1.50 [1.38-1.63] | $7.39 \times 10^{-22}$ | 1.25 [1.13-1.38] | $1.03 \times 10^{-5}$ | 1.54 [1.43-1.67] | $1.13 \times 10^{-27}$ | 1.26 [1.15-1.38] | $4.86 \times 10^{-7}$ |
| **Always** | 1.22 [1.12-1.33] | $2.04 \times 10^{-6}$ | 1.21 [1.11-1.32] | $1.66 \times 10^{-5}$ | 1.16 [1.02-1.33] | 0.03 | 1.15 [1.00-1.32] | 0.05 | 1.48 [1.36-1.61] | $5.16 \times 10^{-20}$ | 1.27 [1.14-1.42] | $8.87 \times 10^{-6}$ | 1.72 [1.60-1.85] | $1.40 \times 10^{-50}$ | 1.37 [1.24-1.50] | $7.44 \times 10^{-11}$ |
| **Females** | | | | | | | | | | | | | | | | |
| **Sometimes** | 1.04 [0.96-1.13] | 0.32 | 0.99 [0.91-1.08] | 0.88 | 0.96 [0.86-1.07] | 0.46 | 0.95 [0.84-1.06] | 0.34 | 1.23 [1.17-1.29] | $3.80 \times 10^{-16}$ | 1.19 [1.12-1.26] | $3.41 \times 10^{-9}$ | 1.17 [1.11-1.23] | $2.00 \times 10^{-9}$ | 1.08 [1.02-1.14] | $4.71 \times 10^{-3}$ |
| **Usually** | 1.36 [1.20-1.55] | $1.88 \times 10^{-6}$ | 1.16 [1.02-1.32] | 0.03 | 1.15 [0.94-1.39] | 0.17 | 1.10 [0.90-1.34] | 0.37 | 1.25 [1.14-1.36] | $6.70 \times 10^{-7}$ | 1.24 [1.12-1.37] | $2.64 \times 10^{-5}$ | 1.22 [1.14-1.30] | $8.38 \times 10^{-10}$ | 1.06 [0.99-1.14] | 0.09 |
| **Always** | 1.12 [1.03-1.21] | $4.82 \times 10^{-3}$ | 1.01 [0.93-1.10] | 0.85 | 1.23 [1.07-1.41] | $3.29 \times 10^{-3}$ | 1.04 [0.90-1.19] | 0.63 | 1.32 [1.21-1.44] | $1.85 \times 10^{-10}$ | 1.35 [1.21-1.49] | $1.89 \times 10^{-8}$ | 1.32 [1.25-1.39] | $4.33 \times 10^{-22}$ | 1.05 [0.98-1.13] | 0.19 |

*Odds ratios are in comparison to the "Never" group.*

*The fully adjusted model is adjusted for age, sex, BMI, TDI and other employment factors.*

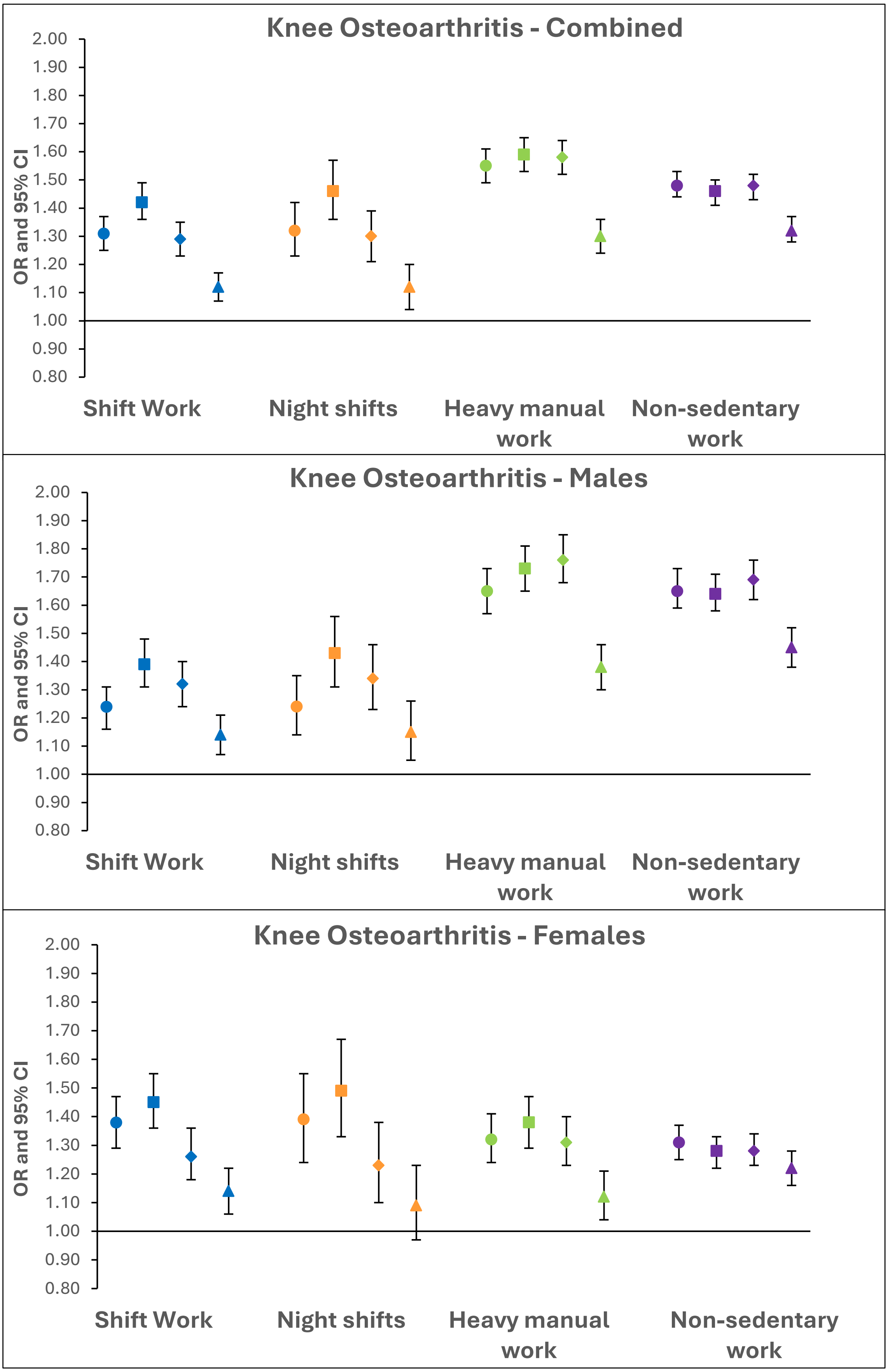


***Supplementary Figure 1 – Logistic regression results for the associations between binary work exposures and knee osteoarthritis in combined and sex-stratified analyses.***

*Odds ratios with 95% confidence intervals displayed. Different shapes represent the various adjustments - circle: unadjusted (model 1), square: age and sex (model 2), diamond: age, sex, BMI and TDI (model 3) and triangle: age, sex, BMI, TDI and other work variables (model 4).*

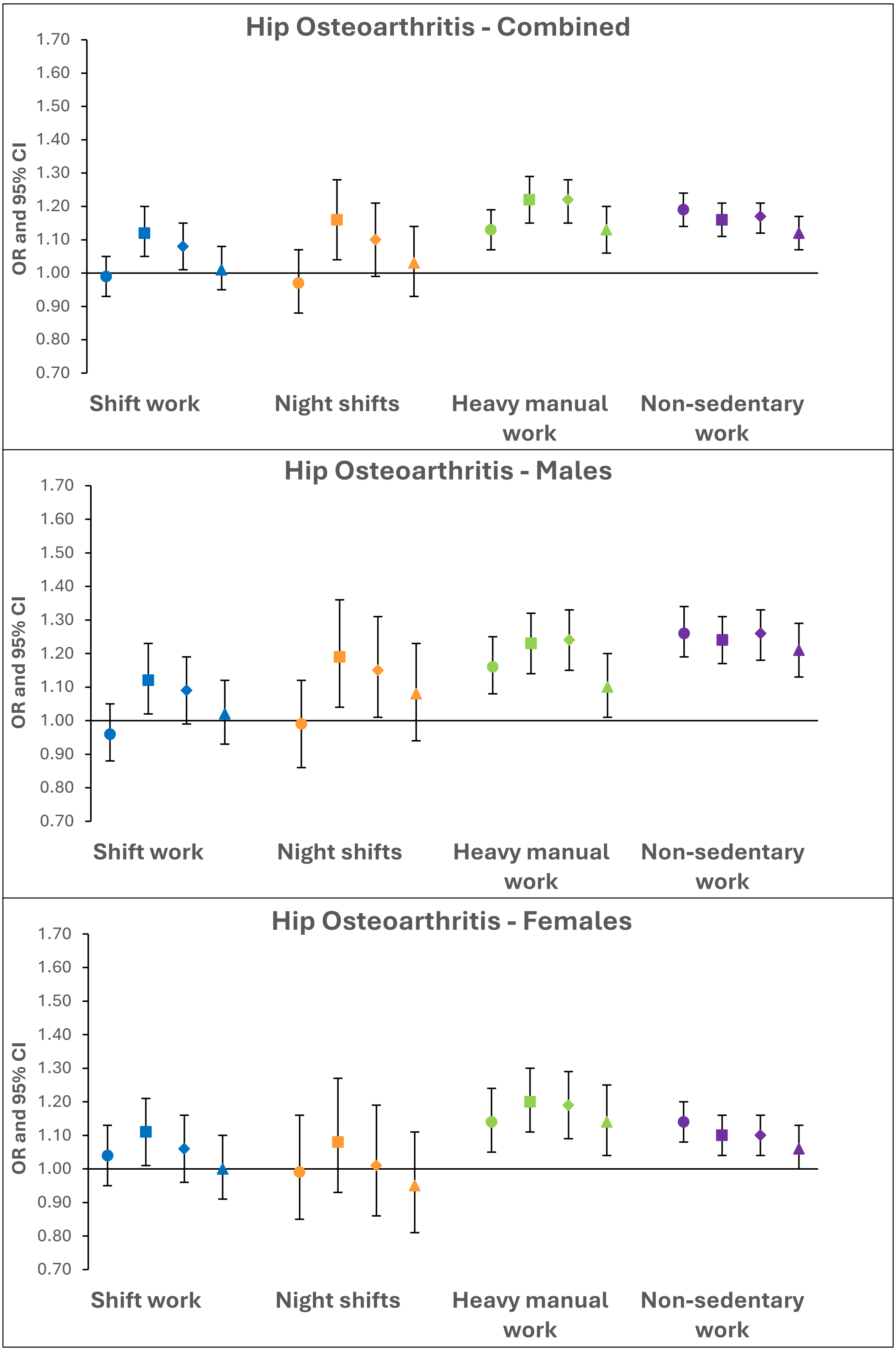


***Supplementary Figure 2 – Logistic regression results for the associations between binary work exposures and hip osteoarthritis in combined and sex-stratified analyses.***

*Odds ratios with 95% confidence intervals displayed. Different shapes represent the various adjustments - circle: unadjusted (model 1), square: age and sex (model 2), diamond: age, sex, BMI and TDI (model 3) and triangle: age, sex, BMI, TDI and other work variables (model 4).*

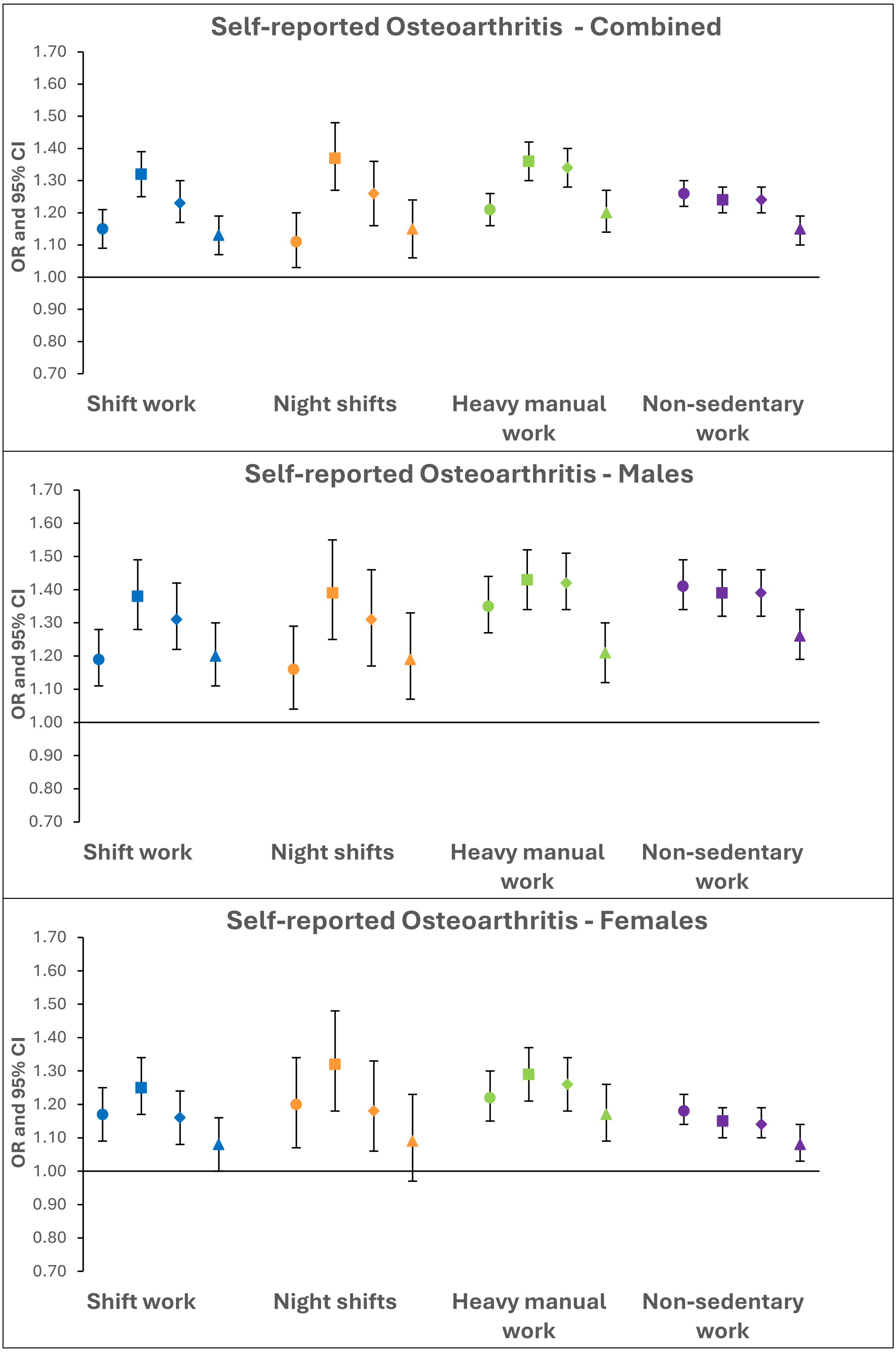


***Supplementary Figure 3 – Logistic regression results for the associations between binary work exposures and self-reported osteoarthritis in combined and sex-stratified analyses.***

*Odds ratios with 95% confidence intervals displayed. Different shapes represent the various adjustments - circle: unadjusted (model 1), square: age and sex (model 2), diamond: age, sex, BMI and TDI (model 3) and triangle: age, sex, BMI, TDI and other work variables (model 4).*
